# Acceptability and Feasibility of a Multicomponent Intervention to Improve Acute Myocardial Infarction Care in Northern Tanzania: the MIMIC Pilot Trial

**DOI:** 10.1101/2024.12.13.24319026

**Authors:** Julian T. Hertz, Francis M. Sakita, Kelvin F. Haukila, Pankrasi S. Shayo, Frida M. Shayo, Joyce Willy, Godfrey Lameck, Emmanuel Kisanga, Hayden B. Bosworth, Janet P. Bettger, Faraan O. Rahim

## Abstract

**Background:** The Multicomponent Intervention to Improve Acute Myocardial Infarction Care (MIMIC) was developed to increase uptake of evidence-based care for acute myocardial infarction in Tanzania. MIMIC consists of five components: triage cards, pocket cards, an online training module, patient educational pamphlets, and clinical champions. Our aim was to determine the acceptability and feasibility of this intervention among emergency department (ED) providers in Tanzania.

**Methods:** During a one-year pilot of the MIMIC intervention at the Kilimanjaro Christian Medical Centre in northern Tanzania, ED physicians and nurses were approached and invited to complete a survey eliciting their perspectives on MIMIC. The survey included the four-item Acceptability of Intervention Measurement (AIM) and four-item Feasibility of Intervention Measurement (FIM) tools. Mean AIM and FIM scores were generated by assigning scores of 1-5 for each response (1= strongly disagree, 2 = disagree, 3= neutral, 4= agree, 5= strongly agree), and dividing by four.

**Results:** Sixty-four participants were enrolled, including 27 (42%) physicians and 37 (58%) nurses. The mean AIM score was 4.82 (sd = 0.31) out of a maximum possible score of 5. The mean FIM score was 4.61 (sd 0.47). Of participants, 63 (98%) reported using the pocket cards and 54 (84%) reported completing the training module, which took a mean of 16.5 (sd 13.3) minutes to complete. Of 36 nurses who worked in triage, all (100%) reported using the MIMIC triage cards.

**Conclusions:** The MIMIC intervention is highly acceptable and feasible in a northern Tanzanian ED. Use of a co-design approach in the development of the MIMIC intervention likely increased the acceptability and feasibility the intervention to staff. Additional study is needed to determine the effectiveness of this intervention on clinical care processes and patient outcomes.

**Trial Registration:** ClinicalTrials.gov NCT04563546; registered on September 21^st^, 2020; https://clinicaltrials.gov/study/NCT04563546

## Introduction

Acute myocardial infarction (AMI) is a life-threatening emergency and a leading contributor to morbidity and mortality worldwide.[1] In sub-Saharan Africa (SSA), the rising incidence of cardiovascular risk factors such as hypertension and obesity has significantly increased the burden of AMI throughout the region.[2] Epidemiological studies estimate that the AMI burden in SSA has increased by 71.4% since 1990,[3] and AMI contributes to over a quarter of a million deaths each year in the region.[4] In northern Tanzania, recent evidence suggests that the burden of AMI is particularly large: the local incidence of AMI is similar to the United States, and AMI is common among patients presenting to Tanzanian EDs with chest pain.[5–7]

Despite these trends, studies from Tanzania have shown that uptake of evidence-based AMI care is sub-optimal in the country. In the ED of a referral hospital in northern Tanzania, we found that only half of adult patients with possible AMI symptoms underwent electrocardiogram (ECG) or cardiac biomarker testing, and approximately 90% of AMI cases were missed in routine ED care.[6, 8] Additionally, less than 25% of AMI patients were given aspirin or evidence-based therapies, and few continued appropriate preventative treatments post-hospitalization, leading to a staggering 43% thirty-day mortality rate for AMI in the region, one of the highest reported globally.[6, 8, 9]

To address these gaps in AMI care, a team of local emergency physicians partnered with international experts in implementation science, emergency medicine, and cardiology to develop the Multicomponent Intervention to Improve Acute Myocardial Infarction Care in Tanzania (MIMIC).[10] Adapted from the ACS-BRIDGE intervention implemented in Brazil,[11] MIMIC leverages rigorous implementation science methods to address system-related, provider-related, and patient-related barriers to AMI care.[10, 12] However, the acceptability and feasibility of the MIMIC intervention have yet to be assessed. Therefore, a pilot trial of the MIMIC intervention was conducted at Kilimanjaro Christian Medical Centre (KCMC) in northern Tanzania. In this manuscript, we report the acceptability and feasibility of the MIMIC intervention, which were the primary implementation outcomes of this pilot trial. The results of this study will elucidate the potential utility of MIMIC in improving the uptake of evidence-based AMI care in Tanzania and will inform future trials of the intervention elsewhere in SSA.

## Methods

### Study Setting

The study was conducted at KCMC, the same tertiary care center where our preliminary studies identifying gaps in evidence-based AMI care were conducted.[6, 8, 9, 13] Located in the urban center of Moshi, Tanzania, KCMC provides healthcare services to over 15 million people throughout the northern regions of the country. The facility has an ED that receives patients 24 hours per day and is staffed by a team of physicians, clinical officers, and nurses. The ED is stocked with basic AMI diagnostic tools, including ECGs and both point-of-care and laboratory-based troponin assays. It is also equipped with basic AMI medications, such as aspirin, clopidogrel, heparin, statins, nitrates, beta-blockers, and thrombolytics. While KCMC offers outpatient follow-up appointments for AMI patients, the facility currently lacks cardiac specialists and the capacity for percutaneous coronary intervention. The KCMC ED is staffed by 70 full-time staff, including physicians, clinical officers, and nurses; members of the KCMC staff are fluent in both English and Swahili.

### Study Procedures

#### MIMIC Implementation

MIMIC was implemented from September 1st, 2023, to August 31st, 2024, by KCMC ED staff; the research team was not involved in implementation. The MIMIC quality improvement intervention consists of five key components, described in detail elsewhere.[10] First, patients presenting to the KCMC ED with chest pain or dyspnea were identified by triage nurses, and their stretchers were affixed with “AMI Suspect” cards that prompted ED clinicians to consider AMI diagnosis and follow corresponding protocols for care. Second, all ED physicians were provided pocket cards to serve as an accessible reference on MI diagnosis and treatment procedures. Third, patients who were diagnosed with AMI were provided an educational pamphlet on AMI management, including information on healthy diet, physical activity, and smoking cessation. Fourth, ED physicians and nurses were required to complete a one-time online module reviewing evidence-based AMI diagnosis and treatment. Fifth, an ED physician and nurse designated as “AMI champions” were tasked with monitoring compliance with MIMIC components, rewarding clinicians for outstanding AMI care with congratulatory certificates, and auditing AMI care in the KCMC ED.

#### Participant Selection & Eligibility Criteria

Implementation outcomes were assessed via surveys administered to KCMC ED staff, including ED physicians, clinical officers, and nurses. ED staff were enrolled daily from 8 AM to 11 PM, seven days a week, throughout the MIMIC pilot trial. All KCMC ED clinicians and administrative staff members working during the study period were eligible for participation in the surveys. Written, informed consent was required from all participants during enrollment. No further exclusion criteria restricted study participation. To minimize social desirability bias, participants self-administered the electronic survey on a tablet. A research assistant sat with the participant during survey completion to answer any questions, but the research assistant did not view the participant’s responses.

#### Recruitment Procedures

A convenience sampling approach was used: ED staff were approached directly at KCMC and offered participation by trained research assistants. Surveys were conducted during break periods, meals, or shift conclusions to minimize disruptions to clinical care or work environment. Each staff member was compensated 5,000 TSH (approximately 2 USD) for their time completing their survey.

#### Provider Surveys

Implementation outcomes were assessed via surveys to KCMC ED providers (**Supplementary Material**). Provider surveys were conducted from one month after the initiation of MIMIC (October 1^st^, 2023) until the conclusion of the pilot trial (September 1^st^, 2024). These surveys included the four-item Acceptability of Intervention Measure (AIM)[14] and the four-item Feasibility of Intervention Measure (FIM)[14] to evaluate provider perceptions of MIMIC acceptability and MIMIC feasibility, respectively. Additional survey questions assessed provider attitudes towards individual MIMIC components, as well as supplemental measures of feasibility, such as the amount of time providers spent completing the online module. Survey questions were written in both English and Swahili; the survey was translated and back-translated from English to Swahili to ensure accuracy and comprehensibility of all survey items.

### Implementation Outcomes Definitions

#### Acceptability

Acceptability, the primary outcome of this pilot trial, was measured via the four-item AIM instrument embedded within the provider survey.[14] The response to each AIM question is measured on a 5-point Likert Scale, and a numerical score of 1-5 was assigned to each response (Strongly Agree=5, Agree=4, Neutral=3, Disagree=2, Strongly Disagree=1). The responses to each question were averaged to give each respondent’s mean AIM score, and the average of all participant scores was calculated for the overall mean acceptability score. An overall mean AIM score ≥4 was defined *a priori* to indicate MIMIC’s acceptability.[15]

#### Feasibility

Feasibility was primarily assessed via the FIM instrument embedded within the provider surveys.[14] Responses to the FIM questions are also measured using a 5-point Likert Scale, and the same process of averaging responses described above was used to evaluate the overall mean feasibility score. A score ≥4 was determined *a priori* to indicate MIMIC’s feasibility. Supplemental evaluations of feasibility were performed via additional questions in the survey, including the reported amount of time ED clinicians needed to complete the online module and the proportion of staff reporting losing their pocket cards.

#### Sample size and analytic approach

The primary study outcome was the acceptability of the MIMIC intervention, as measured by the AIM instrument. In order to obtain a maximally representative sample, we sought to survey at least 90% of the ED staff (at least 63 of the 70 staff members). All statistical analyses were performed in the R Suite (Ver 4.2.1). Simple descriptive statistics were used to present the survey results: categorical variables are presented as proportions and continuous variables are presented as means with standard deviations.

#### Ethics and Data Availability

Ethical approval for this study was obtained from the Tanzania National Institute for Medical Research (NIMR/HQ/R.8a/Vol. IX/2436, Ver 7, Feb 23^rd^, 2021), Kilimanjaro Christian Medical Centre (Proposal 893, Ver 7, Dec 21^st^, 2020), and Duke Health (Pro00090902, Ver 1.7, Sep 16^th^, 2020). The protocol for the pilot trial was registered in ClinicalTrials.gov (NCT04563546). Written, informed consent was obtained from all research participants at enrollment via study forms that were available in both English and Swahili. Study data are available from the corresponding author upon reasonable request.

## Results

During the study period, 64 ED staff members were approached for study participation, all of whom (100%) consented to participate. Study participant characteristics are summarized in Table 1. Slightly over half of participants (n=37, 58%) were nurses; the remainder were physicians.

**Table 1.** Characteristics of Study Participants (N=64)

|  | n | % |
| --- | --- | --- |
| Gender |  |  |
| Male | 41 | 64% |
| Female | 23 | 36% |
| Age, years, mean (sd) | 29.6 (5.7) |  |
| Provider type |  |  |
| Specialist physician<sup>a</sup> | 1 | 2% |
| General physician | 16 | 25% |
| Intern physician | 10 | 16% |
| Nurse | 33 | 52% |
| Intern nurse | 4 | 6% |

<sup>a</sup> Residency trained in Emergency Medicine

Participant responses to AIM items are presented in Table 2. Most participants (≥ 80%) strongly agreed with each AIM item, and no participant disagreed with any of the items. The mean AIM score was 4.82 (sd=0.31) out of a maximum possible score of 5.

**Table 2.** Acceptability of the MIMIC intervention among ED providers at KCMC, 2023-2024 (N= )

| Item | Strongly<br>disagree,<br>n (%) | Disagree,<br>n (%) | Neutral/<br>unsure,<br>n (%) | Agree,<br>n (%) | Strongly<br>agree,<br>n (%) |
| --- | --- | --- | --- | --- | --- |
| The MIMIC intervention meets my approval | 0 (0%) | 0 (0%) | 0 (0%) | 8<br>(13%) | 56<br>(88%) |
| The MIMIC intervention is appealing to me | 0 (0%) | 0 (0%) | 1 (2%) | 12<br>(19%) | 51<br>(80%) |
| I like the MIMIC intervention | 0 (0%) | 0 (0%) | 0 (0%) | 10<br>(16%) | 54<br>(84%) |
| I welcome the MIMIC intervention | 0 (0%) | 0 (0%) | 0 (0%) | 13<br>(20%) | 51<br>(80%) |

Participant responses to FIM items are presented in Table 3. Most respondents (≥91%) agreed or strongly agreed with each FIM item. The mean FIM score was 4.61 (sd 0.47) out of a maximum possible score of 5.

**Table 3.** Perceived feasibility of the MIMIC intervention among ED providers at KCMC, 2023-2024 (N= )

| Item | Strongly<br>disagree,<br>n (%) | Disagree,<br>n (%) | Neutral/<br>unsure,<br>n (%) | Agree,<br>n (%) | Strongly<br>agree,<br>n (%) |
| --- | --- | --- | --- | --- | --- |
| The MIMIC intervention seems<br>implementable | 1 (2%) | 0 (0%) | 0 (0%) | 23<br>(36%) | 40<br>(63%) |
| The MIMIC intervention seems<br>possible | 0 (0%) | 0 (0%) | 0 (0%) | 18<br>(28%) | 46<br>(72%) |
| The MIMIC intervention seems<br>doable | 0 (0%) | 0 (0%) | 0 (0%) | 23<br>(36%) | 41<br>(64%) |
| The MIMIC intervention seems easy<br>to use | 0 (0%) | 0 (0%) | 6 (9%) | 21<br>(33%) | 37<br>(58%) |

Table 4 presents responses to supplemental questions exploring the feasibility of the MIMIC intervention. All but one participant reported receiving and using their pocket card. Approximately one-fourth of participants had to get a replacement pocket card due to loss or damage. Of participants, 57 (89%) reported being instructed to complete the online training module and 54 (84%) actually completed it. Participants spent a mean of 16.5 minutes completing the training module. All nurses who worked in triage reported using the red AMI triage cards.

**Table 4.** Responses to supplemental questions regarding feasibility of the MIMIC intervention among ED providers at KCMC, 2023-2024 (N=64)

| Item | n | (%) |
| --- | --- | --- |
| Did you receive a pocket card? |  |  |
| Yes | 63 | 98% |
| No | 1 | 2% |
| Did you use the pocket card? |  |  |
| Yes | 63 | 98% |
| No | 0 | 0% |
| Didn't receive one | 1 | 2% |
| Did you ever lose your pocket card? |  |  |
| Yes | 12 | 19% |
| No | 52 | 81% |
| Did you have to get a replacement pocket card because yours was lost or damaged? |  |  |
| Yes | 18 | 28% |
| No | 47 | 73% |
| Were you told to complete the online training module? |  |  |
| Yes | 57 | 89% |
| No | 7 | 11% |
| Did you complete the online training module? |  |  |
| Yes | 54 | 84% |
| No | 10 | 16% |
| Why didn't you complete the only training module? (N=10)<sup>a</sup> |  |  |
| I was not told to complete it | 4 | (6%) |
| I forgot | 3 | (5%) |
| I did not have time | 1 | (2%) |
| I did not think it would be helpful | 1 | (2%) |
| Technical difficulties | 1 | (2%) |
| Minutes spent completing the online training module, mean (sd)<sup>b</sup> | 16.5 (13.3) |  |
| As a triage nurse, did you use the red AMI triage cards for patients with possible AMI symptoms? (N=36)<sup>c</sup> |  |  |
| Yes | 36 | (100%) |
| No | 0 | (0%) |

<sup>a</sup> Among 10 providers who reported not completing the training module
<sup>b</sup> Among 54 providers who reported completing the online training module
<sup>c</sup> Among 36 nurses who worked in triage

Table 5 presents the acceptability of individual MIMIC components among survey respondents. Overall, each component of the intervention was well-liked by participants, with 100% of participants reported that they liked or strongly liked each component. The most popular component of the intervention overall was the special red AMI triage cards, which were strongly liked by 57 (89%) of participants. Although only 54 (89%) participants completed the online training module, the vast majority of those who completed the module (n=47, 87%) strongly liked the module.

**Table 5.** Acceptability of individual components of the MIMIC intervention among ED providers at KCMC, 2023-2024 (N= )

| Item | Strongly<br>disagree,<br>n (%) | Disagree,<br>n (%) | Neutral/<br>unsure,<br>n (%) | Agree,<br>n (%) | Strongly<br>agree,<br>n (%) |
| --- | --- | --- | --- | --- | --- |
| Overall, I liked the pocket cards | 0 (0%) | 0 (0%) | 0 (0%) | 14<br>(22%) | 50<br>(78%) |
| Overall, I liked the online training<br>module<sup>a</sup> | 0 (0%) | 0 (0%) | 0 (0%) | 7<br>(13%) | 47<br>(87%) |
| Overall, I liked the system of using<br>special red triage cards for patients<br>with possible AMI symptoms | 0 (0%) | 0 (0%) | 0 (0%) | 7 (11%) | 57<br>(89%) |
| Overall, I liked having a dedicated<br>physician and nurse champion for MI<br>care quality improvement | 0 (0%) | 0 (0%) | 0 (0%) | 10<br>(16%) | 54<br>(84%) |
| Overall, I liked the educational<br>pamphlets for patients | 0 (0%) | 0 (0%) | 0 (0%) | 8<br>(13%) | 56<br>(88%) |

<sup>a</sup>Among 54 providers who completed the online training module

## Discussion

To our knowledge, this is the first study to evaluate a quality improvement intervention for MI care in SSA. We found that the MIMIC intervention, which was developed for the Tanzanian context, was highly acceptable and highly feasible. ED providers reported strongly positive attitudes towards individual components of the intervention and most providers reported engaging with the intervention as it was designed. The MIMIC intervention was developed via a co-design approach, in which the intended users (Tanzanian ED staff) were involved in every step of intervention development and refinement;[10, 16] this approach likely contributed to the acceptability and feasibility of the final intervention in a Tanzanian ED. Although these results are promising, additional study is needed to determine the impact of the intervention on clinical outcomes, such as uptake of evidence-based therapies and mortality.

The overall mean AIM score in our study was 4.82, substantially higher than our pre-determined threshold for acceptability. Respondents reported strongly positive feelings towards each of the five MIMIC components, particularly the red AMI triage cards and the patient educational pamphlets. There were no components of MIMIC for any respondent reported negative feelings. Although the AIM instrument is a well-validated method of determining acceptability of an intervention,[14] additional qualitative study is needed to explore the reasons for acceptability as well as potential ways to make the intervention even more acceptable to ED providers in Tanzania. To our knowledge, there are no other published interventions to improving AMI care in SSA; given the high levels of acceptability observed in our study, the MIMIC intervention may serve as a useful starting point for teams elsewhere in SSA developing their own quality improvement programs.

The overall mean FIM score in our study was 4.61, which was also higher than our pre-determined threshold for feasibility. This result is reinforced by our supplemental measures of feasibility. Specifically, most respondents reported receiving and using the pocket cards, suggesting that this low-cost strategy was highly feasible. Moreover, only a minority of respondents reported losing or having to replace their pocket cards. Similarly, most respondents reported completing the training module, using a median of 16.5 minutes to complete the module, which likely would not be substantially burdensome to ED providers across Tanzania. Notably, 16% of providers did not complete the online training module, primarily because they were not instructed to do so or they forgot. In future implementation studies of MIMIC, additional attention will be needed to ensure all providers are reminded to complete the module. Finally, among nurses working in triage, all reported using the special AMI triage cards, suggesting that this strategy was not overly disruptive to their daily work flow.

This study had several limitations. Firstly, this was a single center study, so the generalizability of our findings is unknown. Additional research will be needed to determine the acceptability of the MIMIC intervention elsewhere in Tanzania. Secondly, the AIM and FIM instruments were provided in both English and Swahili to survey participants, but the psychometric properties of the Swahili versions of these instruments are unknown. Additional research is needed to validate these instruments in Swahili, which will be particularly necessary for study populations that have limited English literacy. Thirdly, as with any survey, our results are subject to social desirability bias; participants may have been inclined to report positive attitudes towards the intervention if they perceived this was desirable to the research team. In order to minimize the bias, participants self-administered the survey and their responses were not visible to any member of the research team in any identifiable way.

## Conclusions

In conclusion, in northern Tanzania, an intervention to improve AMI care was highly feasible and acceptable to ED staff. Given its acceptability and feasibility in our setting, the MIMIC intervention should be considered for adaptation by other groups working to improve AMI care in SSA.

## Data Availability

All data produced in the present study are available upon reasonable request to the authors

## Contributions to the literature

- MIMIC is the first intervention developed to improve acute myocardial infarction care in sub-Saharan Africa.
- In a pilot trial of MIMIC at a Tanzanian hospital, the intervention was highly feasible and acceptable among hospital staff.
- Use of a co-design approach in developing the MIMIC intervention likely contributed to its acceptability and feasibility.

## Declarations

### Ethics approval and consent to participate

Ethical approval for this study was obtained from the Tanzania National Institute for Medical Research (NIMR/HQ/R.8a/Vol. IX/2436, Ver 7, Feb 23^rd^, 2021), Kilimanjaro Christian Medical Centre (Proposal 893, Ver 7, Dec 21^st^, 2020), and Duke Health (Pro00090902, Ver 1.7, Sep 16^th^, 2020). All participants provided written informed consent prior to study participation.

### Availability of data and material

Study data and materials are available from the corresponding author upon reasonable request.

### Author Contributions

JTH, HBB, JPB, and FMSakita conceptualized the study. KFH, PSS, JW, GL, and PL conducted the investigations for this study. FOR and JTH curated the data. JTH, FMShayo, and FMSakita supervised the study. JTH and EK conducted the data analysis for the study. JTH obtained funding for the study. FOR and JTH drafted the manuscript. All co-authors edited the manuscript for critical scientific content.

## Acknowledgements

The study authors gratefully acknowledge the staff of the emergency department at Kilimanjaro Christian Medical Centre for their collaboration in this study.

## Consent for publication

Not applicable

